# COVID-19 pandemic scenario in India compared to China and rest of the world: a data driven and model analysis

**DOI:** 10.1101/2020.04.20.20072744

**Authors:** Manisha Mandal, Shyamapada Mandal

## Abstract

The COVID-19 is a rapidly spreading respiratory illness caused with the infection of SARS-CoV-2. The COVID-19 data from India was compared with China and rest of the world. The average values of daily growth rate (DGR), case recovery rate (CRR), case fatality rate (CFR), serial interval (SI) of COVID-19 in India was 17%, 8.25%, and 1.87%, and 5.76 days respectively, as of April 9, 2020. The data driven estimates of basic reproduction number (R_0_), average reproduction number (R) and effective reproduction number (R_e_) were 1.03, 1.73, and 1.35, respectively. The results of exponential and SIR model showed higher estimates of R_0_, R and R_e_. The data driven as well as estimated COVID-19 cases reflect the growing nature of the epidemic in India and world excluding China, whereas the same in China reveal the involved population became infected with the disease and moved into the recovered stage. The epidemic size of India was estimated to be ∼30,284 (as of April 15, 2020 with 12,370 infectious cases) with an estimated end of the epidemic on June 9, 2020. The R_e_ values in India before and after lockdown were 1.62 and 1.37 respectively, with SI 5.52 days and 5.98 days, respectively, as of April 17, 2020, reflecting the effectiveness of lockdown strategies. Beyond April 17, 2020, our estimate of 24,431 COVID-19 infected cases with lockdown is 78% lower compared to the 112,042 case estimates in absence of lockdown, on April 27, 2020. To early end of the COVID-19 epidemic, strong social distancing is important.

## 1. Introduction

The alarmingly contagious global pandemic COVID-19, caused by SARS-CoV-2, started in December 2019 in Wuhan, China [1], and has now spread to almost all countries and, as of April 9, 2020, infected 1,604,252 people globally, including 81,907 cases, with 3,336 deaths, in China and 1,522,345 cases, with 92,378 deaths, in 203 countries/territories/areas outside China [2]. In India, the first COVID-19 case was reported on January 30, 2020, followed by three cases on February 3, 2020, who were students returned from Wuhan University, with further no new cases till March 1, 2020, but with two more positive cases reported on March 2, 2020 [3]. Confirmed cases in India crossed 50 on March 10, 2020, amongst which 39 had an international travel history, and 16 out of the first 44 cases were foreign nationals. The number of confirmed cases increased to 100 on March 14, 2020, and crossed 1,000 on March 29, 2020, and 14,000 on April 17, 2020, with 486 deaths [2]. In order to contain the ongoing COVID-19 pandemic, India announced a 21-day lockdown that commenced on March 25, 2020 [3], and thereafter extended up to May 3, 2020.

In order to control the spread of the virus and to break the chain of transmission, different non-pharmaceutical strategies such as testing and isolation of COVID-19 cases, contact tracing and quarantine combined with social distancing, masking in public and practicing personal protection are of great significance for the epidemic control [4-6]. In addition, exploring the epidemiological features of COVID-19 will help to understand the nature of the disease in context to the Indian scenario of ongoing global COVID-19 pandemic that was first seen in Wuhan of Hubei province, China [1].

Modeling plays a significant role in understanding the basic epidemiology of an infectious disease and in evaluating the effectiveness of implementing control strategies. The current study stands for the exploration of the epidemic condition of COVID-19 in India, compared with the situation in China, which is the primary epicenter of the ongoing COVID-19 pandemic, as well as in countries outside China, utilizing more recent data available up to April 9, 2020. This study, thus, estimates the epidemiologic parameters including basic reproduction number (R_0_), average reproduction number (R) and effective reproduction number (R_e_), serial interval (SI), transmission rate (β), recovery rate (γ) of COVID-19 infection based on publicly available epidemiological data, exponential and SIR (Susceptible-Infectious-Recovered) models on ongoing COVID-19 pandemic, in order to identify the underlying epidemiologic pattern, evaluation of the efficacy of control strategies, and forecast the epidemiologic dynamics.

## 2. Method

### 2.1. Data source

The data, on ongoing COVID-19 pandemic, including the number of cumulative infected, infectious and recovered cases, and deaths, were retrieved from Worldometer, available up to April 9, 2020, for three geographical locations: India, China, and countries outside China [2]. The lockdown effect in India was studied up to April 17, 2020, retrieving data from Worldometer [2].

### 2.2. Exponential model

The short term prediction of pandemic was done using exponential model, by nonlinear least square method for data fitting and parameter estimation [7]. In India, and the countries outside China, initially the pandemic dynamics was exponential with a growth rate λ, such that the size of the infected population at time *t, N*_*t*_ was given by *N*_*t*_=*N*_0_*e*^*λt*^, and *N*_0_, a constant, was estimated from the fitted exponential curve of the cumulative number of infected cases. This exponential equation was used to determine the SI, which was defined as the time period between the onset of symptoms in the infector (a primary COVID-19 case-patient) and onset of symptoms in the infectee (a patient receiving the infection from the infector) [8]. The SI may be same as the generation time (renewal time of the infected population), considering the onset of symptoms to be same as the onset of infectiousness [9]. Determination of SI is directly linked to calculating the reproduction number (R), which is the number of infectees generating from one infector.

The basic reproductive number (R_0_) of COVID-19 is defined as the average number of secondary infections produced by an infected individual in an otherwise susceptible host population [10]. The probability density function (PDF), expressed as 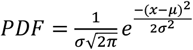, of the serial interval (SI) of COVID-19 was modeled as Weibull distribution with a mean probability density and standard deviation, to determine 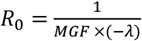, where MGF is the moment generating function of SI.

### 2.3. SIR model

A time-varying SIR model was developed on the basis of the cumulative number of COVID-19 cases per day, and the total population size (N) was divided into three mutually exclusive infection status, assuming that any infectious person (I), has a probability of contacting any susceptible person (S), and later recovered (R), so that *N* =*S* + *I* + *R*. The dynamics of the pandemic in three geographical locations were modeled using the following three differential equations: 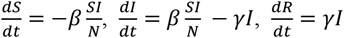 where *β* and *γ* represent transmission rate and recovery rate respectively, defined as the probability of a susceptible-infected contact resulting in a new infection, and the probability of an infected case recovering and moving into the resistant phase, respectively. The reproduction number was calculated using the ratio of the transmission rate to the recovery rate obtained from SIR modeling. The effective reproduction number R_e_, defined as the mean number of secondary cases generated by a primary case at time t in a population, was calculated as an indicator to measure the transmission of COVID-19 both before and after the interventions [10]. The dynamic SIR model was constructed to determine the reproduction numbers, forecast the end date and the final size of the epidemic, by considering the cumulative number of infected cases and the total population number in a country/region as the initial population size *N*, in optimal and optional model, respectively.

## 3. Results and Discussion

The World Health Organization (WHO), on March 11, 2020, declared COVID-19 a pandemic caused with the infection of severe acute respiratory syndrome coronavirus 2 (SARS-CoV-2), a virus that first emerged in the Wuhan city of Hubei province, China, and rapidly spread in every country in the world [11]. As of April 9, 2020, more than 2.3 million COVID-19 infections had been reported in 210 countries and territories around the world [2]. There have been more than 1.59 lakh deaths reported so far, with the United States, UK, France, Italy, and Belgium reporting the highest numbers [2].

In the present study, the dynamic changes of the epidemiologic parameters were taken into account at three different geographical locations: India, China, and countries outside China. The prevailing epidemic scenario in India was compared with the situation worldwide in countries outside China, and China only as depicted in Figure 1, Figure 2 and Figure 3 respectively, in each figure (a) to (d) representing variation in daily growth rate (DGR) with infected case numbers, observed case numbers with predicted numbers, case recovery rate (CRR) with case fatality rate (CFR), and SI with R, respectively. Since the first detection of COVID-19 case in India on January 30, 2020, 6725 number of cumulative infected cases [mean 899.76, 95% Confidence Interval (CI) 444.63 – 1354.89] [Figure 1(a)] have been registered as of April 9, 2020, amongst which the number of cumulative infectious cases were 5863 [mean 802.13, 95% CI 398.85 – 1205.41] [Figure 1(b)], with a daily growth rate (DGR) ranging from 3.44% to 43.02%, latest 13.67% (mean 16.67%, 95% CI 13.58% – 19.76%) [Figure 1(a)]. The number of recovered patients were 635 [mean 72.44, 95% CI 34.07 to 110.81], with a CRR in the range of 5.40% to 12.19% [mean 8.25%, 95% CI 7.71% to 8.79%] [Figure 1(c)], while 227 deaths were registered [mean 25.18, 95% CI 11.28 to 39.09] with CFR in the range of 1.35% to 3.38% [mean 1.87%, 95% CI 1.52% to 2.23%] [Figure 1(c)].

**Figure 1.**
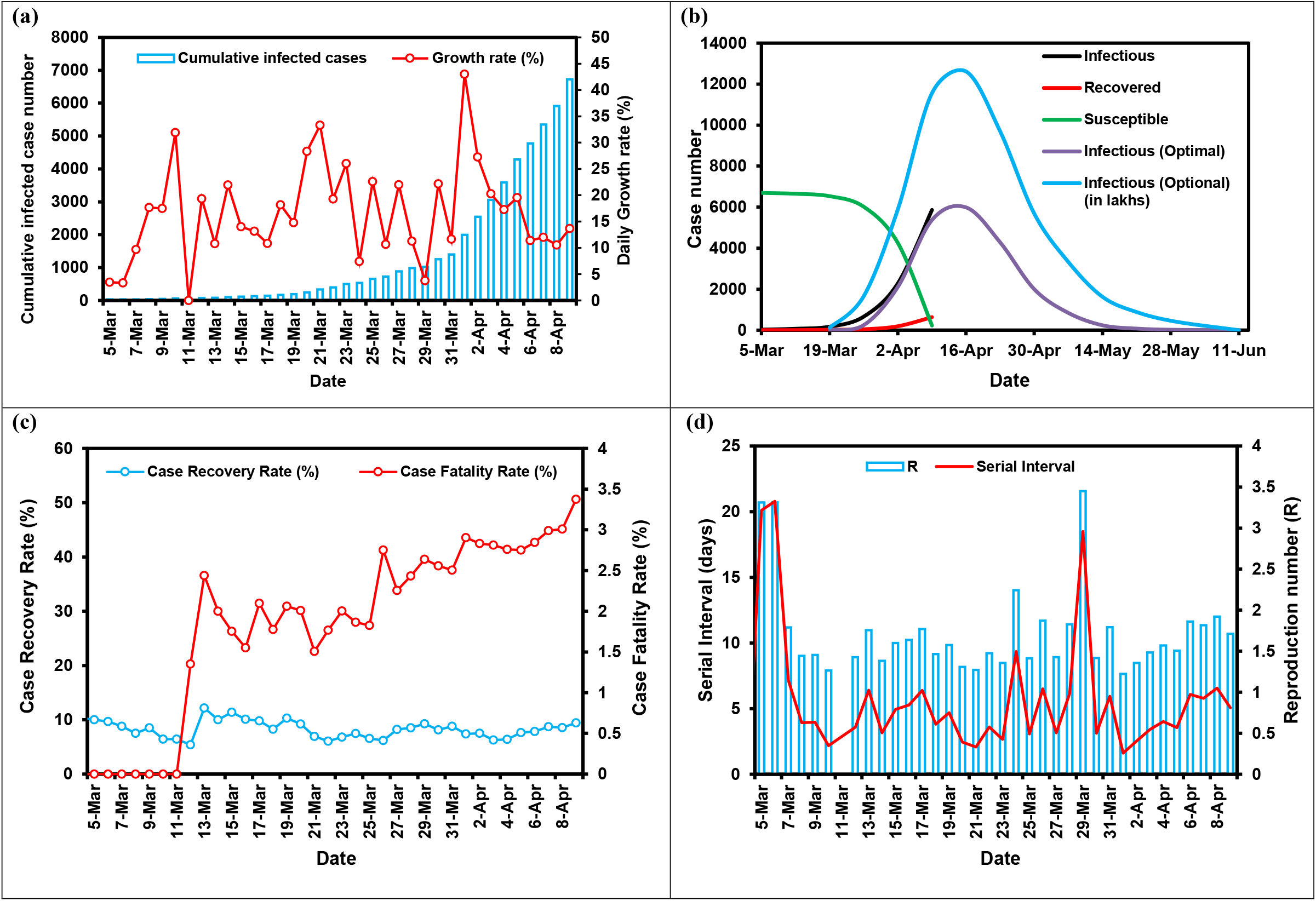
Epidemiological features of COVID-19 pandemic in India: (a) cumulative infected cases and daily growth rate; (b) infectious, recovered and susceptible cases; (c) case recovery rate and case fatality rate; (d) serial interval and reproduction number (R).

**Figure 2.**
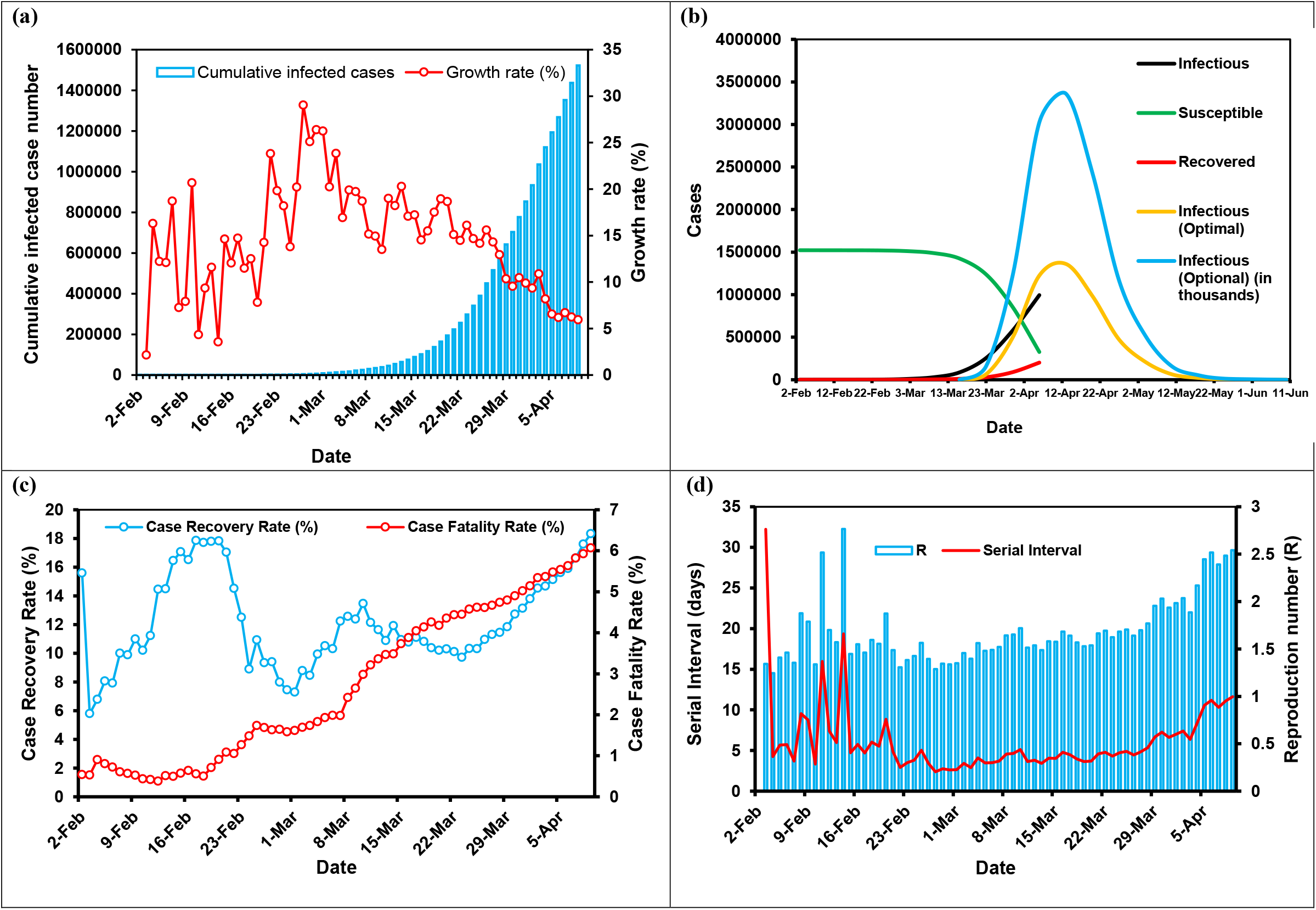
Epidemiological features of COVID-19 pandemic in countries outside China: (a) cumulative infected cases and daily growth rate; (b) infectious, recovered and susceptible cases; (c) case recovery rate and case fatality rate; (d) serial interval and reproduction number (R).

**Figure 3.**
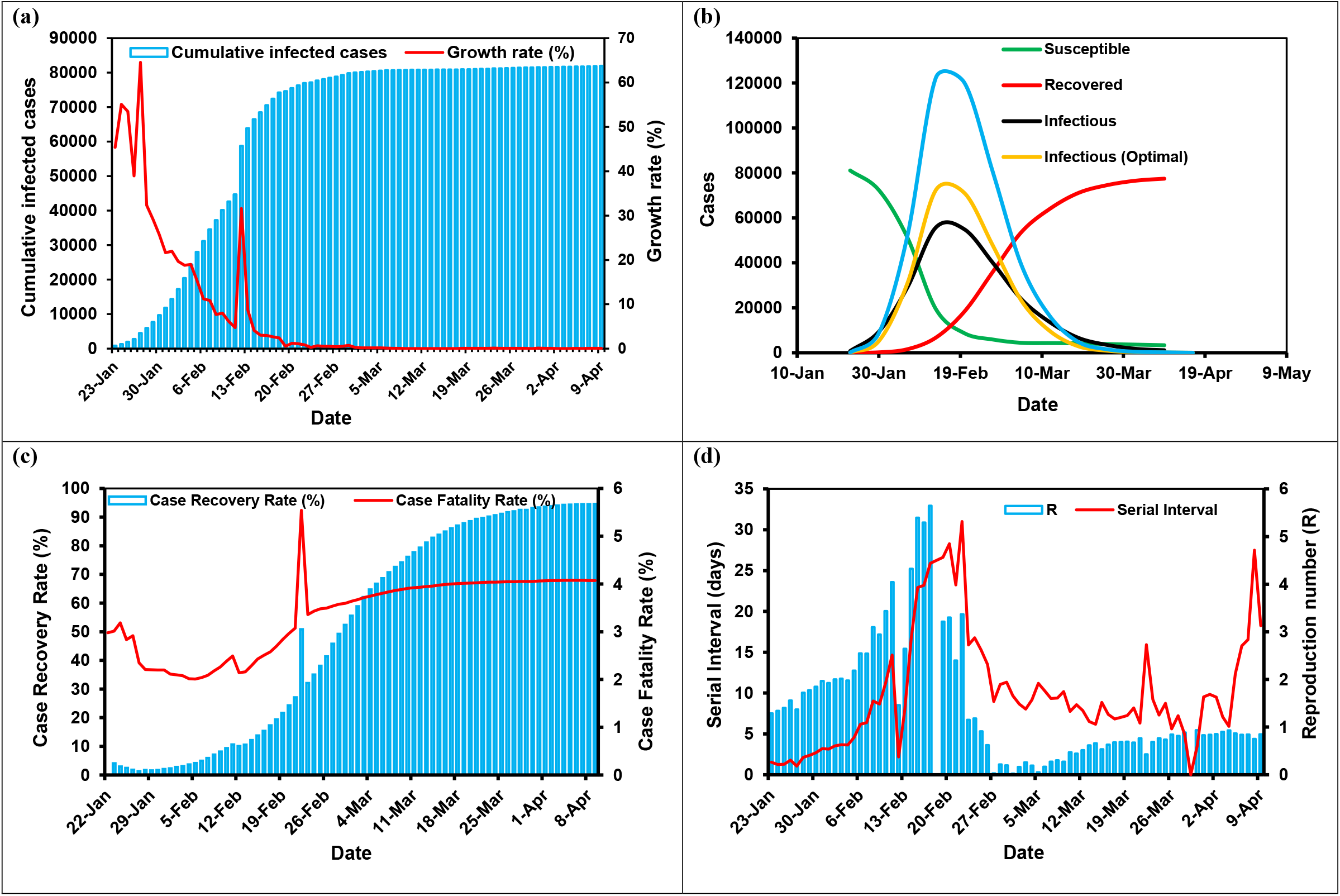
Epidemiological features of COVID-19 pandemic in China: (a) cumulative infected cases and daily growth rate; (b) infectious, recovered and susceptible cases; (c) case recovery rate and case fatality rate; (d) serial interval and reproduction number (R).

An undulating pattern of DGR of COVID-19 infected cases was observed for India as well as world excluding China, but with gradual decrease in the latter for the last few days, while in China the DGR was <0.3 % since February 23, 2020 onwards, as found in the present study. Sanche et al. [12] estimated the growth rate of the early outbreak of COVID-19, in China, as 0.21-0.30 per day. The CFR, which was defined as the ratio of deaths from COVID-19 to the number of cases, provides an assessment of virulence/clinical severity. Therefore, in order to explore the early insights into the severity of ongoing COVID-19 pandemic, CFR of the disease was also estimated. Yang et al. [13], using data-driven statistical method, estimated CFR in the early phase of COVID-19 as 0.15% in mainland China without Hubei, 1.41% in Hubei province without Wuhan city, and 5.25% in Wuhan. The COVID-19 CFR in China has been reported to be as high as 15%, due to low initial case number, which on later time decreased to 3·4% [14]. As of March 17, 2020, in Italy, the CFR of COVID-19 has been recorded as 7.2%, which stands higher than the CFR (2.3%) in China, as of February 11, 2020 [15]; interesting to note, Italy had CFR of 2.3% (as of February 28, 2020), which was identical to the CFR of China [16]. Therefore, though the transmissibility of SARS-CoV-2 is high, the CFR of COVID-19 seems to be lesser compared to that of the other human coronaviruses: SARS (9·5%) and MERS (34·4%), but higher than that of influenza (0·1%) [14]. The CFR in India and the world excluding China showed a rising trend displaying 3.38% and 6.07%, with 49 and 7,233 new deaths, respectively, and an overall death toll of 227 and 92,378, respectively, as of April 9, 2020 [Figure 1 (c), 2 (c)]. The CFR in China reached an equilibrium attaining 4.07 % at the later stage of the COVID-19 epidemic with single new death and 3,336 cumulative deaths as of April 9, 2020 [Figure 3 (c)]. The data driven as well as estimated COVID-19 cases as represented in Figures 1(b) and 2(b), respectively, reflect the ongoing COVID-19 epidemic in India and world excluding China, whereas the same in China [Figure 3(b)] reveal the involved population became infected with the disease and moved into the cured stage.

The SI of COVID-19 infection in India calculated from DGR showed a mean value of 5.44 days, 95% CI of 5.25 to 5.62 days. From the fitted Weibull distribution of the PDF of SI, the mean and standard deviation of 0.67 and of 0.27 was used to obtain MGF value 2.028 to ultimately yield R_0_=3.0 (Table 1). The fitted prediction equation for the size of the infected population at time *t* in India was *N*_*t*_= 0.8057*e*^0.1646*t*^. Applying the exponential model, the fitted prediction equation for the size of the infected population globally at time *t* in countries outside China was *N*_*t*_ = 109.54*e*^0.1497*t*^. The PDF of the SI (mean 6.1 days, 95% CI 5.0 to 7.2 days) of COVID-19 infection worldwide in countries outside China modeled as Weibull distribution showed an average probability density of 0.3151 and standard deviation of 0.0961, MGF value 1.377 to ultimately yield R_0_=4.85 (Table 1). The SI of COVID-19 infection in China, calculated from DGR, showed a mean value of 9.9 days, 95% CI of 8.32 to 11.49 days; which in early phase of the epidemic was <2 days [Figure 3(b)]. The estimate of the average SI as 5.44 days, 6.1 days, and 9.9 days respectively for India, countries outside China, and China as of April 9, 2020, indicate decreasing cycles of COVID-19 transmission from one generation of cases to the next. The SI representing the time from illness onset in a primary case to illness onset in a secondary case is imperative to recognize the turnover of case generations and COVID-19 transmissibility [17]. Sanche et al. [12] determined the doubling time of COVID-19 at early outbreak, in China, as 2.3 – 3.3 days, reflecting the high transmission rate of the virus, SARS-CoV-2. The distribution of COVID-19 SI is a critical input for determining R_0_ and the extent of interventions required to control an epidemic [18]. The mean SI for COVID-19 estimated as of February 8, 2020, by Du et al. [19] was 3.96 days, which was shorter than that found in the current study, and as calculated for SARS (8.4 days) [20], or MERS (14.6 days) [21] as well, implying that contact tracing strategies must contend with the fast replacement of case generations, resulting into number of contacts surpass the existing healthcare facilities [22]. The mean incubation period (time from exposure to symptoms development) for COVID-19 is approximately 5 days (range 1-14 days) [23]. A longer SI than the incubation period is indicative of symptomatic COVID-19 transmission [22], as has been observed for India and world excluding China [Figure 1(d), 2(d)], while China displayed SI in the range of <5 to >5 days implying both pre-symptomatic and symptomatic COVID-19 transmission respectively [Figure 3(d)].

**Table 1:** Reproduction number estimates by different methods.

| Location | Data driven (based on daily growth rate) |  |  | Exponential model* | SIR model |  |  |  |  |  |
| --- | --- | --- | --- | --- | --- | --- | --- | --- | --- | --- |
|  | R<sub>0</sub> | R | R<sub>e</sub> | R<sub>0</sub> | R<sub>0</sub> |  | R |  | R<sub>e</sub> |  |
|  |  |  |  |  | Optimal | Optional | Optimal | Optional | Optimal | Optional |
| India<sup>#</sup> | 1.03 | 1.68 | 1.46 | 3.0 | 1.0 | 1.0 | 5.33 | 3.67 | 4.65 | 3.63 |
| Countries outside China<sup>+</sup> | 1.34 | 1.69 | 1.42 | 4.85 | 7.5 | 7.5 | 9.21 | 7.3 | 7.76 | 7.23 |
| China<sup>++</sup> | 1.29 | 1.4 | 0.36 | – | 7.43 | 7.38 | 3.05 | 0.34 | 0.8 | 0.33 |

\*Based on fitted exponential growth rate and MGF of SI distribution
<sup>#</sup>The optimal and optional SIR model for India was based on the cumulative infected case number = 6,725 on April 9, 2020, and the population of India = $13 \times 10^8$ , as the initial population size, respectively.
<sup>+</sup>The optimal and optional SIR model worldwide for countries outside China was based on the cumulative infected case number = 1,522,345 on April 9, 2020, and the world population excluding China = $37.6 \times 10^8$ , as the initial population size, respectively.
<sup>++</sup>The optimal and optional SIR model for China was based on the cumulative infected case number = 81,907 on April 9, 2020, and the population of China = $13.8 \times 10^8$ , as the initial population size respectively.

The R values derived from DGR showed a variable pattern with time, with increasing trend of R, at recent times, in India (Figure 1d), and world excluding China (Figure 2d). However, the R values based on DGR for China (Figure 3d) showed a stabilized pattern at later stages (R=0.84) as of April 9, 2020, as indicated by the trend of the number of susceptible infections, and recovered cases over time (Figure 3b). Table 1 represents the R_0_, R, and R_e_ values estimated from data based on DGR, exponential and SIR mathematical models for three locations. The estimates of R_0_ and average R produced by SIR mathematical model were higher than that of statistical exponential growth model, and data driven based on DGR (Table 1), possibly attributed to the underlying assumptions of the mathematical models. The earlier studies showed variation of R_0_ that ranged from 1.4 to 6.49 (mean 3.28), for COVID-19 outbreaks, because of variation in sources of data collection, time periods, and modelling methods used [23, 24].

The current estimates depict that the R_0_ values were in the order world(excluding China)> China> India, following DGR as well as SIR methods, while R and R_e_ were in the pattern of world(excluding China)> India> China. This could be explained by the fact that during the initial phase of the epidemic the infected size of COVID-19 was greater in the world, wherein most of the proportion of infected cases were from China, while in the later stage, China contained the SARS-CoV-2 transmission by the time when world excluding China, and India in the later stage, had escalating growth of infected cases. This leads to higher reproduction number compared to that of China with lowest proportion of people susceptible to COVID-19 as well as gradual decrease in the number of cumulative infectious cases (dropping to 1116 as of April 9, 2020), and daily new deaths plummeting trend dropping to one as of April 9, 2020.

The results of the optimal and optional SIR models for India estimate the average R of 5.33 (95% CI 3.05 to 11.33, average β = 0.32, average γ = 0.06), and the average R value of 3.67 (95% CI 2.13 to 7.26, average β = 0.22, average γ = 0.06), respectively, calculated from March 5, 2020 to April 9, 2020, with R_0_ = 1.0. The estimates of the optimal and optional SIR model for countries outside China showed the average R value to be 9.21 (95% CI 8.19 – 10.35) (average β = 0.188, average γ = 0.0204) and the average R value of 7.3 (95% CI 7.24 to 7.26, average β = 0.1491, average γ = 0.0204), respectively, as calculated between February 2, 2020 and April 9, 2020, with R_0_ = 7.5. The optimal and optional SIR model prediction for average R value in China was 3.05 (95% CI 2.89 – 3.22; average β = 0.128, average γ = 0.042) and 0.34 (95% CI 0.33 to 0.35; average β = 0.0143, average γ = 0.042), respectively, as calculated between January 23, 2020 and April 9, 2020, with R_0_ = 7.43 and 7.38 respectively. The reproduction number (R), which determines the transmissibility of the virus, SARS-CoV-2, represents the average number of new COVID-19 cases generated by each of the infected individuals, the initial value of which is regarded as basic reproduction number (R_0_) that depicts the average number of new infections per infected COVID-19 case, while the effective reproduction number (R_e_) has been defined as the average number of secondary cases generated per primary case, with symptom onset, at time t [10]. The R_e_ displays variation as the outbreak progresses in time course and might be affected with control measures. As per the estimate of Zhao et al. [7], R_0_ for SARS-CoV-2 ranged 2.24 – 3.58, which was >1, indicating the potential of the virus to cause and continue outbreaks, and the epidemics to be fade away when the transmissibility could be reduced by 1–1/R_0_ [25].

The epidemic size of India, following optimal SIR modeling, was estimated to be ∼17,886 (as of April 9, 2020 with 5863 infectious cases) and ∼30,284 (as of April 15, 2020 with 12,370 infectious cases) reflecting an estimated end of the epidemic on June 3, 2020, and June 9, 2020, respectively [Figure 1 (b)]. The epidemic size of the world excluding China, following optimal SIR modeling, was estimated to be ∼4,362,777 (as of April 9, 2020 with 1,150,985 infectious cases) and ∼6,303,651 (as of April 15, 2020 with 2,004,136 infectious cases) reflecting an estimated end of the epidemic on June 5, 2020, and June 11, 2020, respectively [Figure 2 (b)]. It has been estimated earlier by Ranjan [26], that R_0_ for India, in relation to COVID-19 pandemic, ranged 1.4-3.9, when data from March 11, 2020 to March 23, 2020 were analyzed, and model within this study predicted an equilibrium of the disease by the end of May, 2020 with infected cases of 13,000 considering the absence of the impact of community transmission and social distancing as well. Currently, China has contained the local transmission of SARS-CoV-2 with unprecedented public health interventions through 76-day massive lockdowns (since January 23, 2020), and that have begun to lift in response to the slowing of the outbreak, which might be informative to public health policy making in other countries [24].

The association of reproduction number with other epidemiological parameters is shown in Table 2. There was significant (p<0.05) association of R with time and SI, globally. The other epidemiologic parameters including the number of infectious cases and total deaths were significantly related to R in China, implying these two variables as an index of R in epidemiologic evolutionary dynamics. However, CFRs in our study were insignificantly associated with R, at three geographical locations, irrespective of the epidemiologic stages. The growth rate of deaths per day has emerged as a useful parameter for tracking the evolution of COVID-19 spread in different regions [27].

**Table 2:** Association of reproduction number with other epidemiological parameters.

| Location | Time (t) in days | Serial interval (SI) | Case fatality rate (CFR) | Number of infectious cases (N) | Number of total deaths (D) | Regression equation |
| --- | --- | --- | --- | --- | --- | --- |
| India | 0.02 | $6.1 \times 10^{-34}$ | 0.98 | 0.56 | 0.71 | $R = 0.0086 \times time + 0.1206 \times SI - 0.0007 \times CFR - 4.4 \times 10^{-5} \times N + 0.0007 \times D + 0.8728$ |
| Countries outside China | 0.04 | $6.8 \times 10^{-6}$ | 0.09 | 0.98 | 0.44 | $R = 0.0133 \times time + 0.0334 \times SI - 0.1219 \times CFR + 1.2 \times 10^{-8} \times N + 8.1 \times 10^{-6} \times D + 1.2432$ |
| China | $3 \times 10^{-5}$ | 0.0002 | 0.72 | 0.00038 | $1.3 \times 10^{-6}$ | $R = 0.0623 \times time + 0.070 \times SI - 0.0891 \times CFR + 3.8 \times 10^{-5} \times N - 0.0016 \times D + 1.3440$ |

(Values expressed as p values, p value < 0.05 significant)

Different mathematical models and studies demonstrated the association between reduction of effective reproduction numbers R_e_ (an indicator to measure the transmission of SARS-CoV-2), effectively to <1, and implementation of public health measures amidst COVID-19 pandemic [24, 28]. India’s strategies with massive social distancing: lockdown since March 25, 2020, and traffic restriction combined with mass contact tracing and quarantine, and isolation of infected people are expected to help curbing the transmission of SARS-CoV-2, and this might be relevant to adopt by other countries facing current test-kit shortages [29]. The mean R_e_ value for the pre-lockdown period (before March 25, 2020) was found to be 1.62 (95% CI 1.36 – 1.89), and the mean R_e_ for the post-lockdown period was 1.1 (95% CI 0.84 – 1.37) (Figure 4), this decline of R_e_ indicated the effectiveness of the introduced interventions. As has been reported by Zhao and Chen [30] in China, before January 30, 2020, the R_e_ was >1 (range: 1.2-5.9), while after January 30, 2020, R_e_ was <1 (range: 0.51-0.53), suggesting the effectiveness of non-pharmaceutical control strategies in averting the spread of SARS-CoV-2.

**Figure 4.**
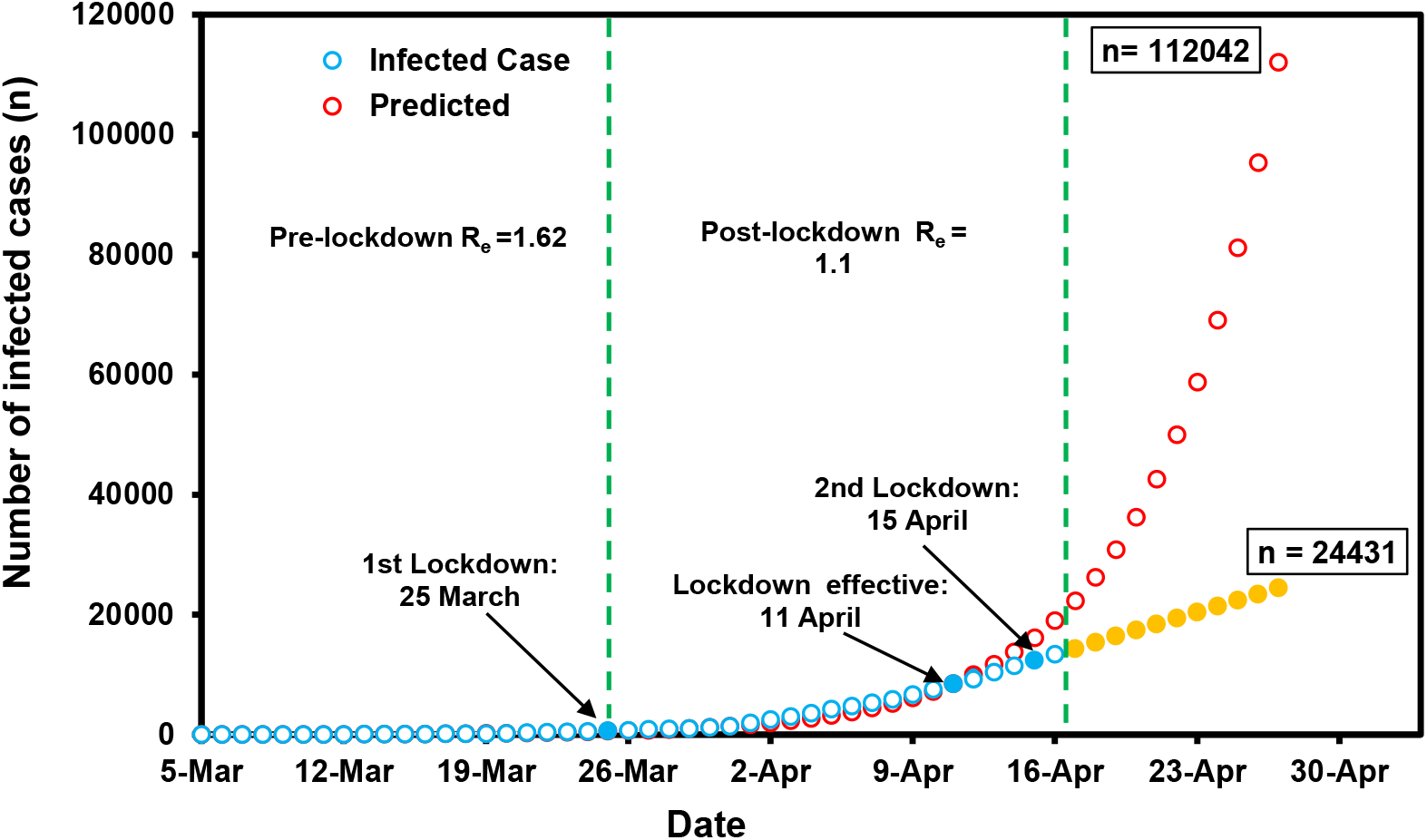
Observed (as on April 17, 2020) and estimated (up to April 27, 2020) infected cases using exponential model. Red hollow circles represent predictions of infected case numbers in India till April 27, 2020 using exponential model. Blue hollow circles represent the observed number of infected cases up to April 17, 2020. Blue filled circles represent the observed number of infected cases and yellow filled circles represent predicted 24,431 number of infected cases in presence of lockdown effect on April 27, which in absence of lockdown represent 1,12,042 cases.

The R_e_ >1 is indicative of escalation of epidemic at time t (unless effective control measures are operated), while R_e_ <1 defines the shrinking of epidemic size at time t, and consequently the outbreak will stop [10]. There has been a gradual decrease of R_e_ from high level of transmission (R_e_: 3.1, 2.6, and 1.9) to <1 (R_e_: 0.9 or 0.5) due to increasing implementation of public health measures [5]. The transmission of SARS-CoV-2 is, however, escalating rapidly worldwide, and multiple countries outside China are now experiencing the devastating consequences of the COVID-19 pandemic that indicates the shortfalls in preparedness. Thus, this is crucial to understand how the stringent strategies (mass lockdown and mass testing alongside) prevented the spread of SARS-CoV-2 in China [31], and to implement this by countries outside China [24].

This study reflected the latest updates about the COVID-19 epidemic up to April 9, 2020. Also besides single R_0_ calculation, the study is an attempt to take into account the dynamic variation of R values on different scenarios of the epidemic wherein the study established the connection between R values and the effect of control interventions in India. Like other countries all over the world, India is experiencing the COVID-19 pandemic since January 30, 2020, and due to its gradual escalation, different states enforced various unprecedented non-pharmaceutical measures, including school and office work closures, alongside testing, contact tracing and quarantine/case isolation, in order to contain the spread of the virus, SARS-CoV-2. Thereafter when the pandemic caused 536 people sick and 10 deaths, India introduced more stringent measures, the countrywide 21-day lockdown enforcement with effect from March 25, 2020, and thereafter, from April 15, 2020, considering the steeping nature of viral transmission in order to stem the disease, COVID-2. The huge physical distancing measure, in the form of country’s complete lockdown, in India, had an astonishing effect in slowing down COVID-19 transmission, and this could be explained with the fact of basic reproduction number (R_0_), the time varying R, and the size of infected population with or without lockdown. The calculated R_0_ =1.03 herein, indicates the capacity SARS-CoV-2 to transmit effectively from one infected patient to more than one susceptible person and to cause epidemic, and thereafter up to March 24, 2020, the average R_e_ value increased to 1.62, and thus, the transmissibility was increased (Figure 4). However, after lockdown the decrease in estimated R_e_ to 1.37 (from March 25, 2020 to April 10, 2020) is an indication of decreased transmissibility, and R_e_=0.46 from April 11, 2020 (the day with 0.44% decrease in predicted infected cases of 8483 to observed infected cases of 8446, in absence and presence of lockdown, respectively, and with susceptible proportion of 0.43) to April 17, 2020 (the day with 36% decrease in predicted infected cases of 22,329 to observed infected cases of 14,352, without and with lockdown, respectively, and with susceptible proportion of 0.034), demonstrating a tremendous effect of lockdown in COVID-19 containment (Figure 4). Beyond April 17, 2020, our estimate of 24,431 COVID-19 infected cases with lockdown is 78% lower compared to the 112,042 case estimates in absence of lockdown, on April 27, 2020 (Figure 4).

## 4. Conclusion

Combined with the available evidences the current findings suggest the adoption of social distancing (countrywide lockdowns, closing schools and workplaces, and restrictive of mass gatherings and travel) measures in order to shrink the peak intensity of outbreaks, wherein the lifting of the measures results resurgence of SARS-CoV-2 transmission. Therefore, high grade effective social distancing strategies help reduce the extent of SARS-CoV-2 infections that cause strain on the health care systems, unless critical care capacity is expanded substantially, or a treatment or vaccine becomes available. Also regular surveillance of SARS-CoV-2 in an urgent manner (even after accomplishing ample control of transmission, if any) is essential for the vigilance of further plausible resurgence of COVID-19 outbreaks. Further, determining CFR of COVID-19 and accounting daily deaths will help track the dynamics of disease spread, formulate control measures and design the supplies of health care systems to mitigate the global crisis of ongoing COVID-19 pandemic. To note that our prediction is of more academic than applied, and the mathematical model estimates (which is based on the publicly available information) will improve as more data will appear due to the novel coronavirus (SARS-CoV-2) infection, and thus, it will be prudent to review the assessments as epidemics develop, if any.

## Data Availability

Basic data were retrieved from publicly available worldometer

## Conflict of Interest

We declare that there is no conflict of interest.

## Funding support

There was no source of funding for this study.

## Notes

### Competing Interest Statement

The authors have declared no competing interest.

### Funding Statement

No funding source for the study

